# Adherence to antiretroviral therapy among Female Sex Workers in Kampala Uganda

**DOI:** 10.1101/2025.03.19.25324247

**Authors:** Denis Max Ssemakula, Sheila N. Balinda, Yunia Mayanja, Onesmus Kamacooko, Andrew Abaasa, Janet Seeley

**Affiliations:** Medical Research Council, UVRI & LSTHM Uganda Research Unit, Plot 51-59, Entebbe, Uganda; London School of Hygiene and Tropical Medicine; UCSF International Traineeships in AIDS Prevention Studies (I-TAPS) program

**Keywords:** Antiretroviral therapy, adherence, female sex workers, information and communication technology, mobile phone

## Abstract

**Background:** Increased access to ART to key populations who bear a disproportionate burden of HIV, including female sex workers, provides a great opportunity to reduce onward transmission. This is however dependent on achieving high levels of adherence to ART. We set out to determine level of adherence to ART and associated factors among female sex workers in Uganda.

**Methods:** In this cross-sectional study between May and June 2017, we consecutively enrolled 226 female sex workers accessing HIV care at an urban clinic in Kampala, Uganda. We assessed self-reported adherence using interviewer administered questionnaires and reviewing medical records. We defined high level adherence as those who scored ≥95% at assessment. Using multivariable logistic regression, we identified factors independently associated with adherence.

**Results:** Overall, 59.2 % of participants were adherent to ART. Major reasons for non-adherence were being away from home (40.8%) and forgetfulness (26.7%). In the multivariable model, owning a phone (AOR 2.90; 95% CI 1.07, 7.88), a 10-year increase in age (AOR 1.60; 95% CI 1.00, 2.60) and being a widow (AOR 0.22; 95% CI 0.05, 0.87) were independently associated with adherence.

**Conclusion:** Our study found low observed adherence to ART among female sex workers in Uganda. This baseline assessment builds a case for the development and scale up of targeted intervention strategies to increase to ART adherence among female sex workers. Incorporating information and communication technology such as mobile phones in routine adherence counselling could be scaled up among key populations such as female sex workers.

## Introduction

The HIV/AIDS remains a public health problem world over, with 38 million infected and 690,000 deaths reported in 2021 [1]. Key populations including and their sexual partners are estimated to have accounted for 62% of new HIV infections globally in 2019 [1]. These include men who have sex with men (MSM), people who inject drugs (PWID) and female sex workers (FSW). Specifically, FSW globally continue to share a disproportionate burden of HIV with an estimated 30 times higher risk of acquiring infection than adults aged 15-49 years in the general population [1]. In sub-Saharan Africa, a systematic review of studies conducted in 25 countries indicated that 36.9% of FSW were HIV positive with HIV prevalence as high as 70.7% [2] in some countries. In Uganda, HIV prevalence among FSW ranges from 32.4 to 52% [3, 4] compared to 9.5% among women in the general population [5]. FSW and their clients are estimated to contribute 18% of the new infections in Uganda [5].

Several factors influence the higher risk of HIV acquisition and thus the high prevalence among FSW. These include alcohol abuse, multiple clients and other sexual partners, inconsistent condom use, and a higher prevalence of sexually transmitted infections [4, 6]. Access to HIV preventive services is limited due to lack of community-based support systems, stigmatisation and criminalisation, high mobility, and the judgemental attitude of some health workers towards FSW [7–10]. Additionally, FSW with high mobility may face additional transport challenges to health facilities [7].

FSW are prioritized for increased access to antiretroviral therapy (ART), to improve their health outcomes and prevent onward transmission of HIV to sex partners (Ref). Since 2013 Uganda started “test and treat” for key populations including FSW [11] which developed into universal ART for all, following WHO guidelines [12]. However, to optimise full benefits of ART, high level adherence sustains viral suppression, reduces risk of drug resistance, and onward HIV transmission [13, 14]. Adherence is defined as the patient’s ability to follow a treatment plan by taking the right medicine, in the right dose, at the right time, and in the right way. Over 95% adherence to ART confers optimal therapeutic effectiveness [15]. Factors that influence adherence to ART include patient-related factors, such as alcohol and other substance abuse [16], age, sex, level of education, stigma, social support and HIV status disclosure [17, 18]. The treatment-related factors are side effects, pill burden, and severity of disease [19]. In addition, factors related to the patient-provider relationship like trust and acceptance [20] and the healthcare system also affect adherence [21]

FSW face unique realities that determine their contact with the health care system. For instance, a study in Kampala found that only 45% of HIV-positive FSW knew their HIV sero-status, of whom only 37.8% reported to be on ART, and only 35.2% achieving viral suppression [22]. Studies have attributed low ART uptake and interrupted engagement in HIV care among FSW to their high mobility [23], an environment of punitive laws and discrimination against FSW, stigma, judgemental attitude from some health workers, frequent arrests, and intimate partner violence [9, 24]. Further, despite policies to increase access to ART among FSW, appreciation of their unique realities of life, and efforts to improve their low agency in the fight against HIV, there is still a paucity of information regarding adherence to ART and its associated factors for FSW in SSA. One systematic review largely from industrialized countries and another from India indicate that adherence to ART in FSW ranges between 48% [25] and 76% [26]. We build on this work with HIV-positive FSW accessing ART from an urban clinic in Kampala Uganda to determine the level of ART adherence and associated factors. Our study assessed adherence to ART and associated factors among HIV-positive FSW attending an urban clinic in Uganda.

## Methods

### Ethical considerations

Our study presented more than minimal risks including potential unintended disclosure of HIV and FSW status to the community during home visits. Noteworthy, we performed a secondary analysis of clinical information without any invasive procedures that diminished some risks. Still, we sought and obtained approved from the Uganda Virus Research Institute Research Ethics Committee (GC/127/14/10/30) and Uganda National Council for Science and Technology (HS 364). Further, we obtained written informed consent from all participants, and all data were de-identified prior to analysis.

### Study design and setting

We conducted a cross-sectional study between May and June 2017. The Good Health for Women Project (GHWP) is in Mengo Kisenyi, a densely populated slum in the central division of Kampala, the capital city of Uganda. The GHWP clinic was established in 2008 by Medical Research Council/Uganda Virus Research Institute (MRC/UVRI) in collaboration with the Uganda Ministry of Health. The GHWP was set up to study the epidemiology of HIV and other sexually transmitted infections (STI) among FSW, and to provide free enhanced HIV prevention services to this key population. These include treatment of common illnesses, comprehensive HIV care, management of other STIs, reproductive health services, condom promotion and distribution, and alcohol risk reduction counselling. These services are delivered at three-monthly visits and according to the clients’ demand. Although GHWP started offering ART in 2013, ‘test and treat’ began in August 2014; a government of Uganda policy where HIV-positive FSW start ART immediately irrespective of CD4 count [11]

### Study participants and eligibility criteria

GHWP patients who are FSW are defined as women who engage in sex with men for money and other favours. FSW are recruited to the clinic at night through a peer-led approach from their operating hotspots (e.g., bars, brothels, night clubs, and streets, eating places, lodges, and guesthouses). Greater detail on the recruitment of FSW to the GHWP clinic are described elsewhere [4]. Eligibility criteria were being 18 years old and above, accessing ART from GHWP and having been on ART for at least 6 months. Participants were enrolled consecutively as they came either for their routine clinic visits or for any other reason.

### ART preparation, initiation, and follow-up

Prior to initiating ART, confirmed HIV-positive clients had baseline CD4 counts done, pre-ART adherence counseling. ART was dispensed first for 14 days, and then one month, two months and three-monthly refills were given only when the client was determined to be stable. A combination of tenofovir, lamivudine and efavirenz (TDF/3TC/EFV) was the first line regimen unless otherwise indicated. For follow up, viral load testing was performed six months after ART initiation, repeated yearly if viral suppression had been achieved and, if not, 4 to 6 monthly following Uganda ministry of health guidelines [27]. Ongoing adherence counseling was provided for the clients at every clinic visit as appropriate. Specifically, those clients for whom viral suppression was not achieved, at least three sessions of intensive adherence counseling were done monthly by the nurse counselor until viral suppression was achieved. Screening for and management of opportunistic infections and other STI was done at every contact with the clients.

### Study variables, measures, and data collection

#### Study outcome

We categorized the outcome variable, ART adherence, as a binary adherence or non-adherent. Adherence was measured using self-reports of missed pills over a thirty-day recall period. Studies have shown that self-reported data correlate with viral load changes [28–30]. Participants who reported taking ≥95% of prescribed pills were considered adherent and those who took <95% of the pills were considered non-adherent.

#### Exposure variables

Potential predictors of ART adherence were categorized into social demographics, those related to disclosure, behavioral variables and treatment related clinical variables. Social demographics variables included age, highest level of education attained, marital status, working in a bar or lodge and owning a phone. Those related to disclosure included: HIV status and ART disclosure to either any significant other or to male regular sexual partner. Social support was defined as a participant having a treatment companion and not living alone. Stigma was defined as the participant never disclosed her HIV status or did not want clinic staff to visit her home for fear of being discriminated by her neighbors. Behavioral variables included mobility and alcohol use in the last three months, as yes/no answers. Mobility was considered as staying at the current location less than 50% of her time or had changed place of residence at least once in the previous one year. Treatment related clinical variables included duration from HIV diagnosis to ART initiation, duration on ART, baseline CD4 count, possession of a medicine reminder tool, possession of an ART adherence card, WHO stage, pregnancy at ART initiation, type of regimen, history of concomitant medications in the past one month and viral suppression defined as viral load of less than 1,000 copies/ml. The most recent test results from the patients’ medical records were preferred to real time testing to meet study costs.

### Data collection and laboratory methods

Our trained study staff collected data using pre-tested interviewer administered tools and data extraction from participant files. Pretesting of the data collection tool was done in the first week of data collection to determine suitability, face validity, comprehension, and to achieve a common understanding of the tool among the nurse counselors. Quality control checks were done for each completed case report form to ensure completeness, accuracy, and correctness.

As part of routine clinical care, blood samples were collected to assess HIV viral load and CD4+ T-cell count. Baseline CD4 + T-cell counts were done on ART initiation and VL were done at least six months from ART initiation. Baseline CD4 + T-cell counts were performed on plasma using Multi-test Trucount tube CD4/CD8/CD3-Lyse no wash (Becton D Biosciences, USA). Viral load testing was performed on plasma using an Abbott Real-time HIV-1 PCR assay (Abbott Park, Illinois, USA) from the Uganda Central Public Health Laboratories

### Statistical analyses

We entered data into epi-data, cleaned and exported to STATA 15.0 (StataCorp, College Station, TX, USA) for analysis. We categorized and summarized participants’ characteristics by counts and percentages. We analyzed continuous variables using appropriate measures of central tendency, means or medians, and their relevant measures of dispersion, standard deviations, and interquartile ranges respectively. We determined the proportion of non-adherent FSW who scored ≥95% at assessment divided by the total number included in the analysis and expressed as a percentage. We used logistic regression to identify factors associated with adherence at bivariate analysis. We selected variables with statistical significance p<0.1 on log likelihood ratio tests (LRT) for multivariable logistic regression. We maintained ‘alcohol use’ variable in the final model since literature shows it is strong predictor of ART adherence. We presented results adjusted odds ratios (AOR) with p-values and 95% confidence intervals (CI). P-values <0.05 were considered significant.

## Results

A total of 5,157 FSW had been cumulatively recruited into the Good Health for Women Project (GHWP), by the end of June 2017 (Figure 1). About two in five, 38.5% (n=1,984) were HIV positive of whom 42.1% (835) accessed ART from the GHWP clinic. The remainder (n=1,149) had either not yet started ART or were receiving ART elsewhere. Among those accessing ART from GHWP, 512 participants (61.3%) were on ART for more than 6 months at GHWP and were eligible for the study, of whom 226 (44.1%) were included in the final analysis of this study. Noteworthy, we excluded the 38.7% (323) on ART for less than six months.

**Figure 1.**
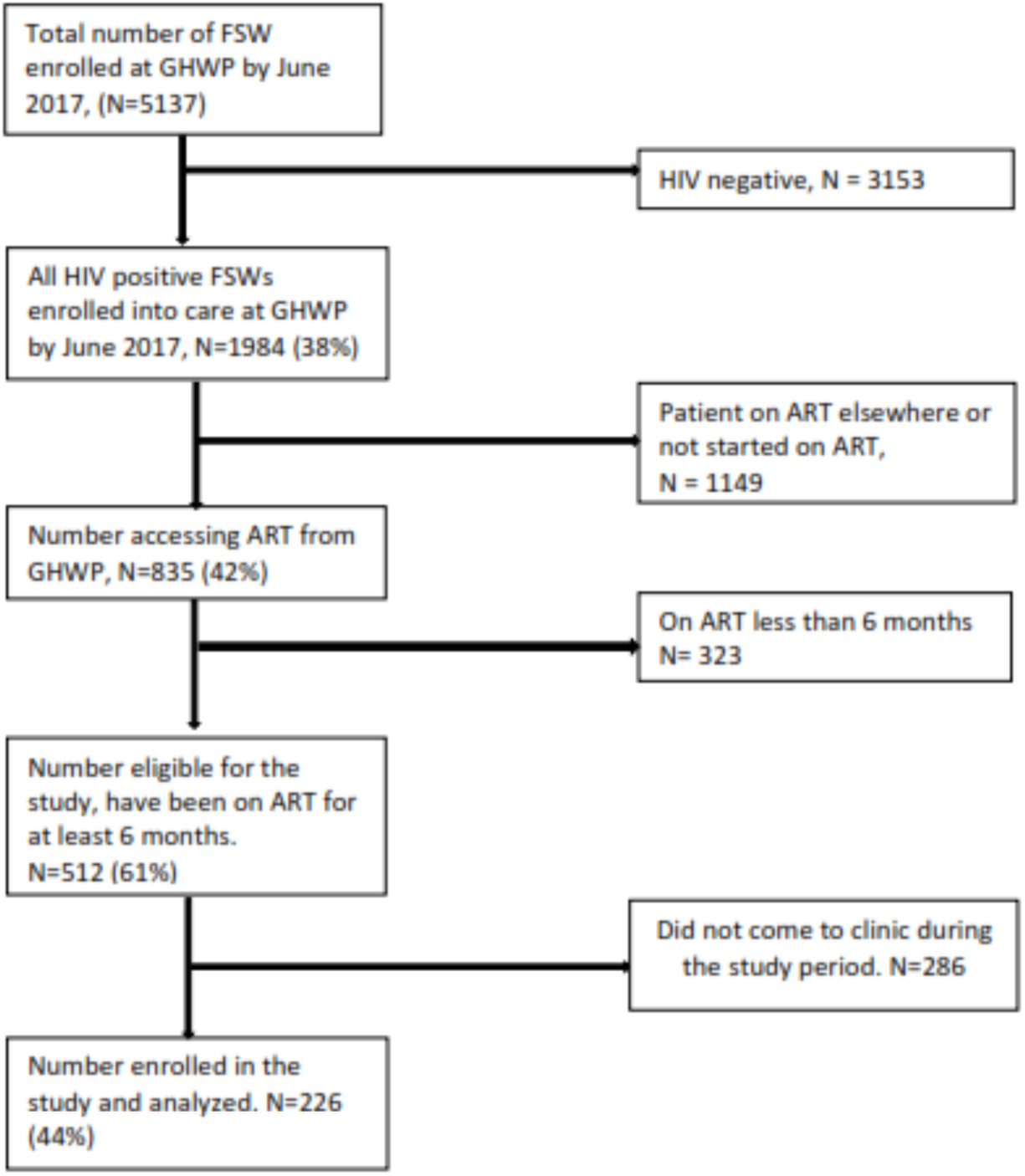
Flow diagram of female sex workers recruited for a study of ART adherence, Kampala, Uganda, 2017.

### Characteristics of study participants

FSW had a mean age of study participants was 32 years (SD±6.5), with 56.2% in the 25–35 age bracket. 56.6% had attained at least primary level education. While only 37.6% were separated or divorced, most FSW (88.5%) reported disclosing their HIV status to a significant other (Table 1). Over half (56.2%) reported consuming alcohol over the last three months. The majority (90.3%) reported owning a mobile phone.

**Table 1:**
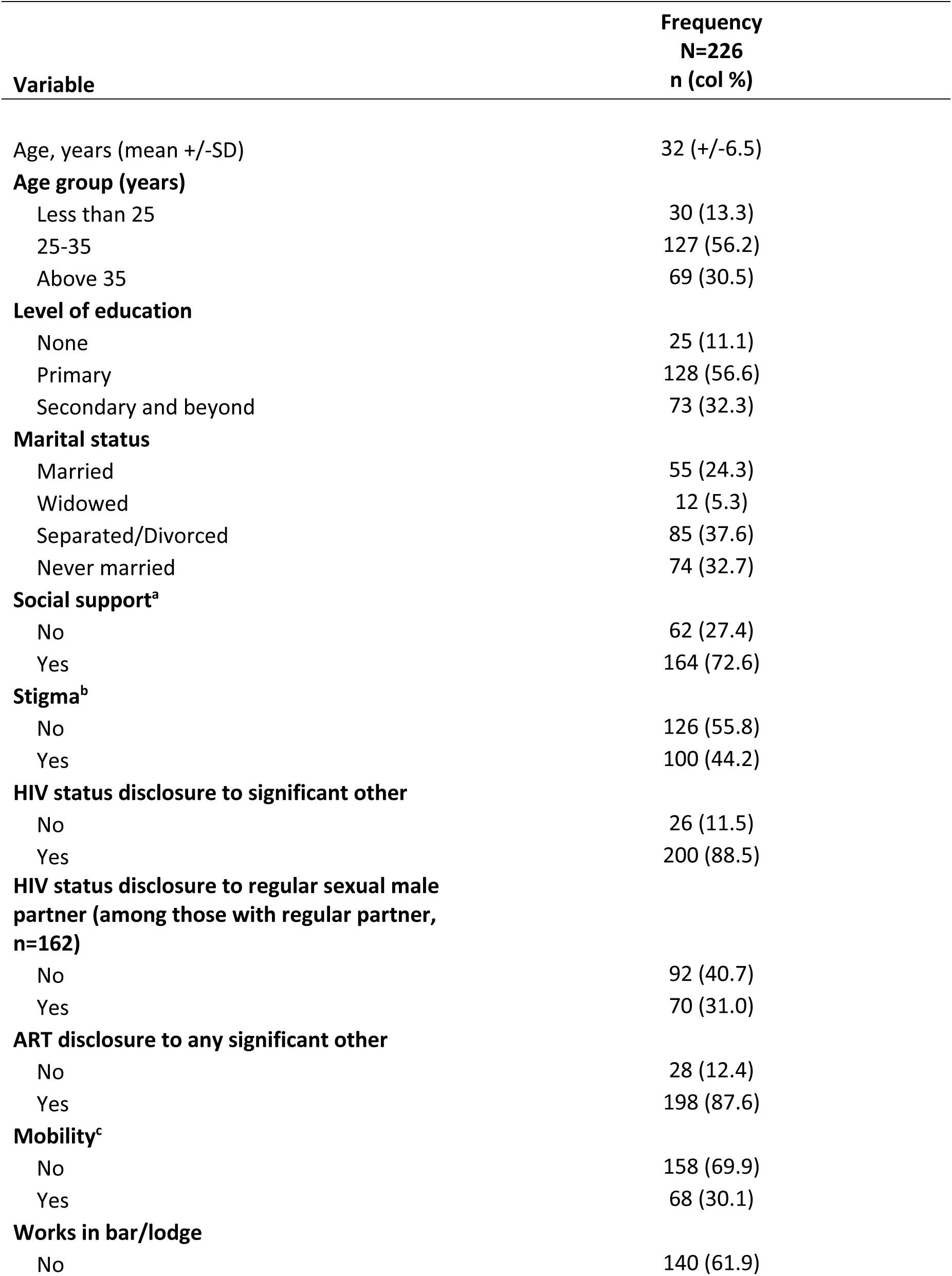
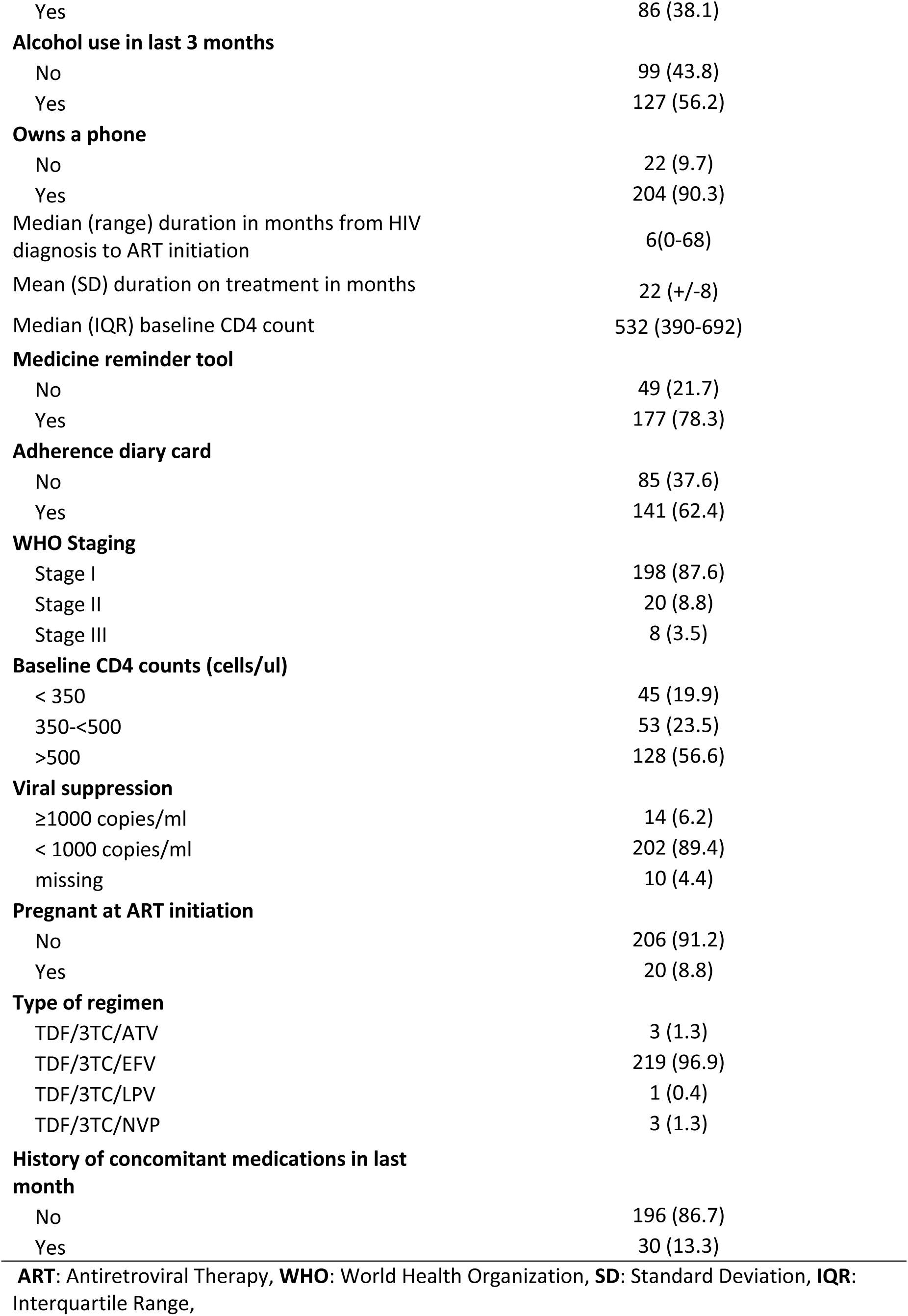
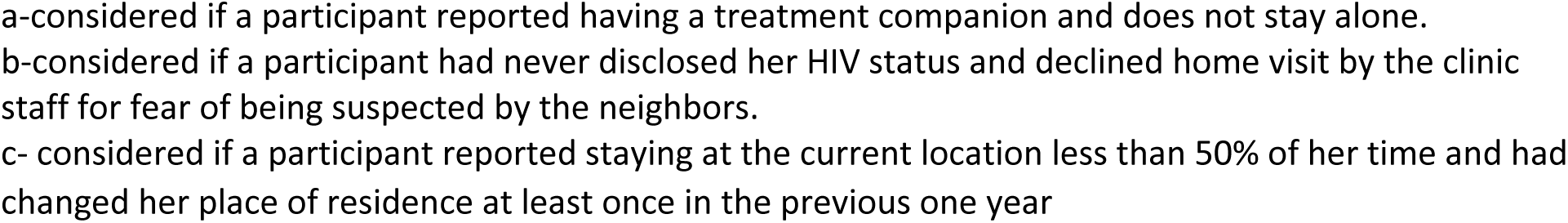
Baseline characteristics of a sample of HIV positive female sex workers (N=226) attending clinic at GHWP, Kampala, Uganda assessed for adherence to ART between the months of May and June 2017.

**Table 2.**
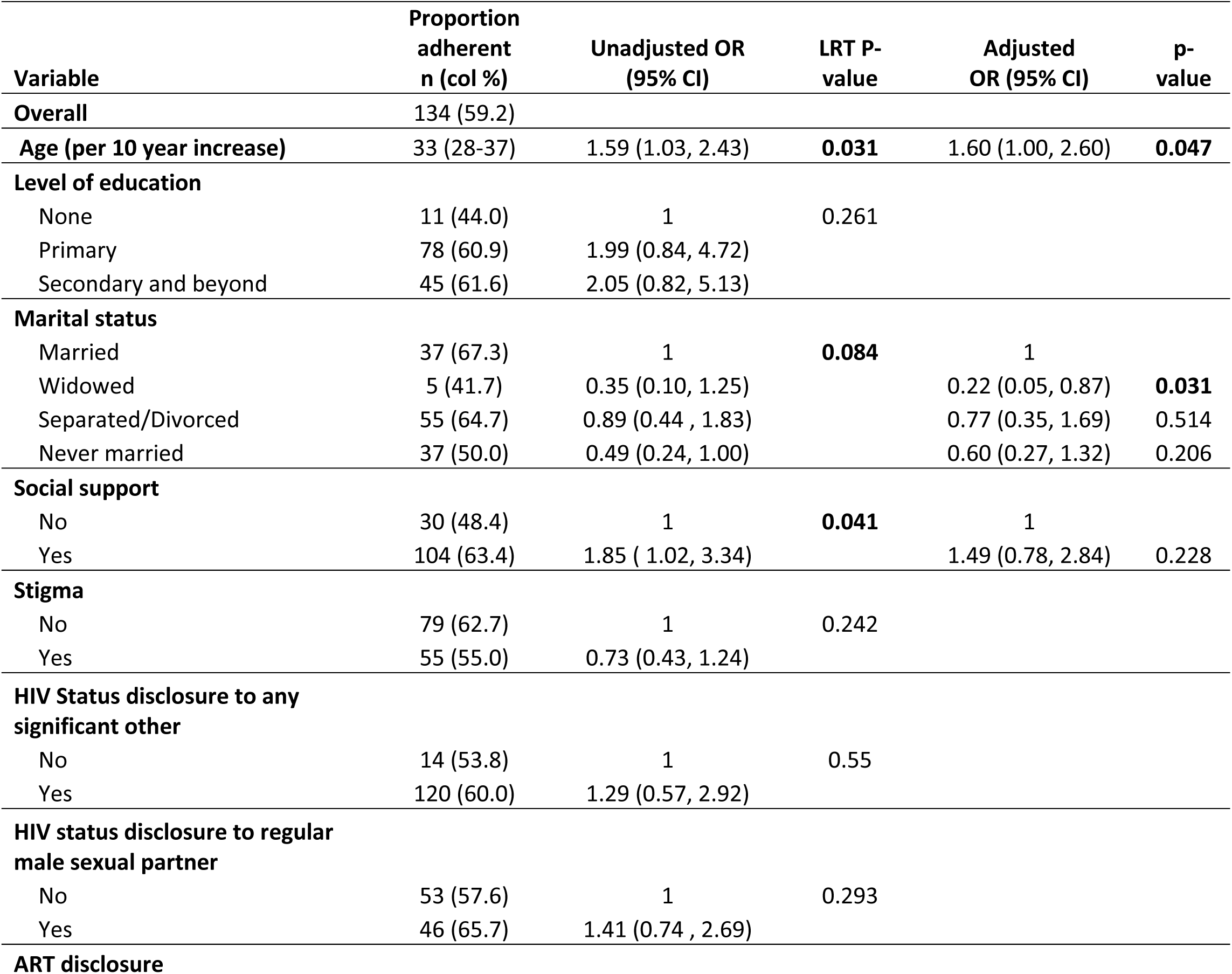
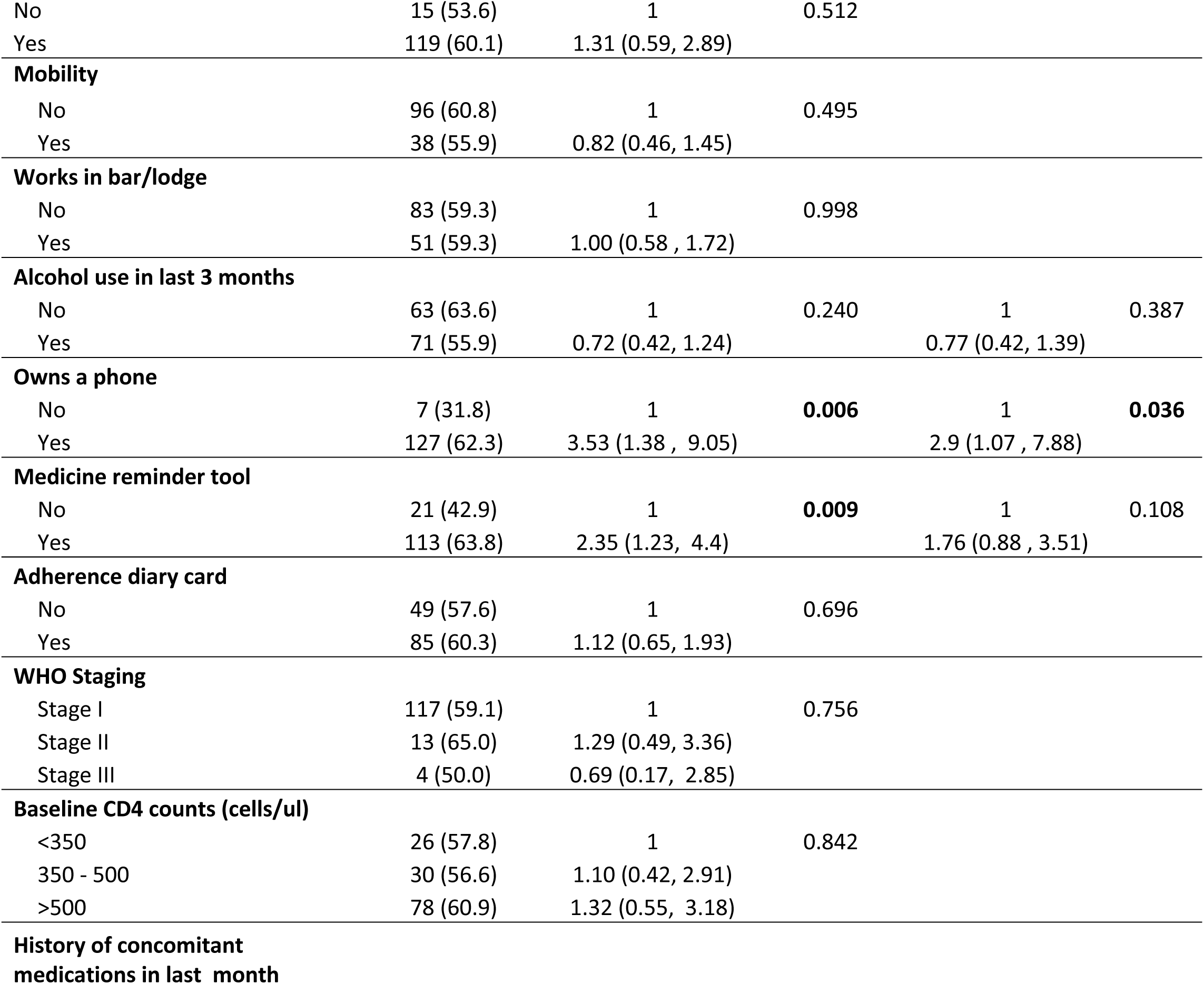
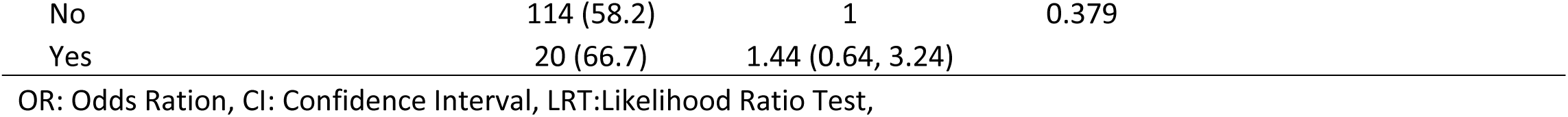
Multivariable regression of adherence to ART and associated factors among HIV-positive female sex workers attending clinic at GHWP, Kampala, Uganda between May and June 2017 (N=226)

| Variable | Proportion<br>adherent<br>n (col %) | Unadjusted OR<br>(95% CI) | LRT P-value | Adjusted<br>OR (95% CI) | p-<br>value |
| --- | --- | --- | --- | --- | --- |
| **Overall** | 134 (59.2) |  |  |  |  |
| **Age (per 10 year increase)** | 33 (28-37) | 1.59 (1.03, 2.43) | **0.031** | 1.60 (1.00, 2.60) | **0.047** |
| **Level of education** |  |  |  |  |  |
| None | 11 (44.0) | 1 | 0.261 |  |  |
| Primary | 78 (60.9) | 1.99 (0.84, 4.72) |  |  |  |
| Secondary and beyond | 45 (61.6) | 2.05 (0.82, 5.13) |  |  |  |
| **Marital status** |  |  |  |  |  |
| Married | 37 (67.3) | 1 | **0.084** | 1 |  |
| Widowed | 5 (41.7) | 0.35 (0.10, 1.25) |  | 0.22 (0.05, 0.87) | **0.031** |
| Separated/Divorced | 55 (64.7) | 0.89 (0.44 , 1.83) |  | 0.77 (0.35, 1.69) | 0.514 |
| Never married | 37 (50.0) | 0.49 (0.24, 1.00) |  | 0.60 (0.27, 1.32) | 0.206 |
| **Social support** |  |  |  |  |  |
| No | 30 (48.4) | 1 | **0.041** | 1 |  |
| Yes | 104 (63.4) | 1.85 ( 1.02, 3.34) |  | 1.49 (0.78, 2.84) | 0.228 |
| **Stigma** |  |  |  |  |  |
| No | 79 (62.7) | 1 | 0.242 |  |  |
| Yes | 55 (55.0) | 0.73 (0.43, 1.24) |  |  |  |
| **HIV Status disclosure to any**<br>**significant other** |  |  |  |  |  |
| No | 14 (53.8) | 1 | 0.55 |  |  |
| Yes | 120 (60.0) | 1.29 (0.57, 2.92) |  |  |  |
| **HIV status disclosure to regular**<br>**male sexual partner** |  |  |  |  |  |
| No | 53 (57.6) | 1 | 0.293 |  |  |
| Yes | 46 (65.7) | 1.41 (0.74 , 2.69) |  |  |  |
| **ART disclosure** |  |  |  |  |  |
| No | 15 (53.6) | 1 | 0.512 |  |  |
| Yes | 119 (60.1) | 1.31 (0.59, 2.89) |  |  |  |
| **Mobility** |  |  |  |  |  |
| No | 96 (60.8) | 1 | 0.495 |  |  |
| Yes | 38 (55.9) | 0.82 (0.46, 1.45) |  |  |  |
| **Works in bar/lodge** |  |  |  |  |  |
| No | 83 (59.3) | 1 | 0.998 |  |  |
| Yes | 51 (59.3) | 1.00 (0.58 , 1.72) |  |  |  |
| **Alcohol use in last 3 months** |  |  |  |  |  |
| No | 63 (63.6) | 1 | 0.240 | 1 | 0.387 |
| Yes | 71 (55.9) | 0.72 (0.42, 1.24) |  | 0.77 (0.42, 1.39) |  |
| **Owns a phone** |  |  |  |  |  |
| No | 7 (31.8) | 1 | **0.006** | 1 | **0.036** |
| Yes | 127 (62.3) | 3.53 (1.38 , 9.05) |  | 2.9 (1.07 , 7.88) |  |
| **Medicine reminder tool** |  |  |  |  |  |
| No | 21 (42.9) | 1 | **0.009** | 1 | 0.108 |
| Yes | 113 (63.8) | 2.35 (1.23, 4.4) |  | 1.76 (0.88 , 3.51) |  |
| **Adherence diary card** |  |  |  |  |  |
| No | 49 (57.6) | 1 | 0.696 |  |  |
| Yes | 85 (60.3) | 1.12 (0.65, 1.93) |  |  |  |
| **WHO Staging** |  |  |  |  |  |
| Stage I | 117 (59.1) | 1 | 0.756 |  |  |
| Stage II | 13 (65.0) | 1.29 (0.49, 3.36) |  |  |  |
| Stage III | 4 (50.0) | 0.69 (0.17, 2.85) |  |  |  |
| **Baseline CD4 counts (cells/ul)** |  |  |  |  |  |
| <350 | 26 (57.8) | 1 | 0.842 |  |  |
| 350 - 500 | 30 (56.6) | 1.10 (0.42, 2.91) |  |  |  |
| >500 | 78 (60.9) | 1.32 (0.55, 3.18) |  |  |  |
| **History of concomitant medications in last month** |  |  |  |  |  |

|  |  |  |  |
| --- | --- | --- | --- |
| No | 114 (58.2) | 1 | 0.379 |
| Yes | 20 (66.7) | 1.44 (0.64, 3.24) |  |

OR: Odds Ration, CI: Confidence Interval, LRT:Likelihood Ratio Test,

Clinically, most FSW (87.6%) had initiated ART early at WHO stage 1 and had a mean duration on ART of 22 months (SD±8). Half FSW had a baseline CD4+ T-cell count above 532 cells/mm^3^(IQR, 390–692). The majority (96.9%) of participants were on first line regimen (TDF/3TC/EFV). Viral suppression (<1,000 copies/ml) was observed in 89.4% of participants.

### Adherence and associated factors

Overall, 59.2 % of participants were adherent to ART. Being away from home was the most important reason for missing pills, 40.8% (n=120) (figure 2). In the bivariate analysis, adherence to ART was more likely among participants who had social support compared to those who did not (OR 1.85; 95% CI 1.02, 3.34; p=0.041), those who owned a phone at the time of the study (OR 3.53; 95% CI 1.38, 9.05; p=0.006), and those who reported possession of a medicine reminder tool (OR 2.35; 95% CI 1.23, 4.40; p=0.009). A 10-year increase in age of the client was associated with increased odds of adherence (OR 1.59; 95% CI 1.03, 2.43; p=0.031).

**Figure 2.**
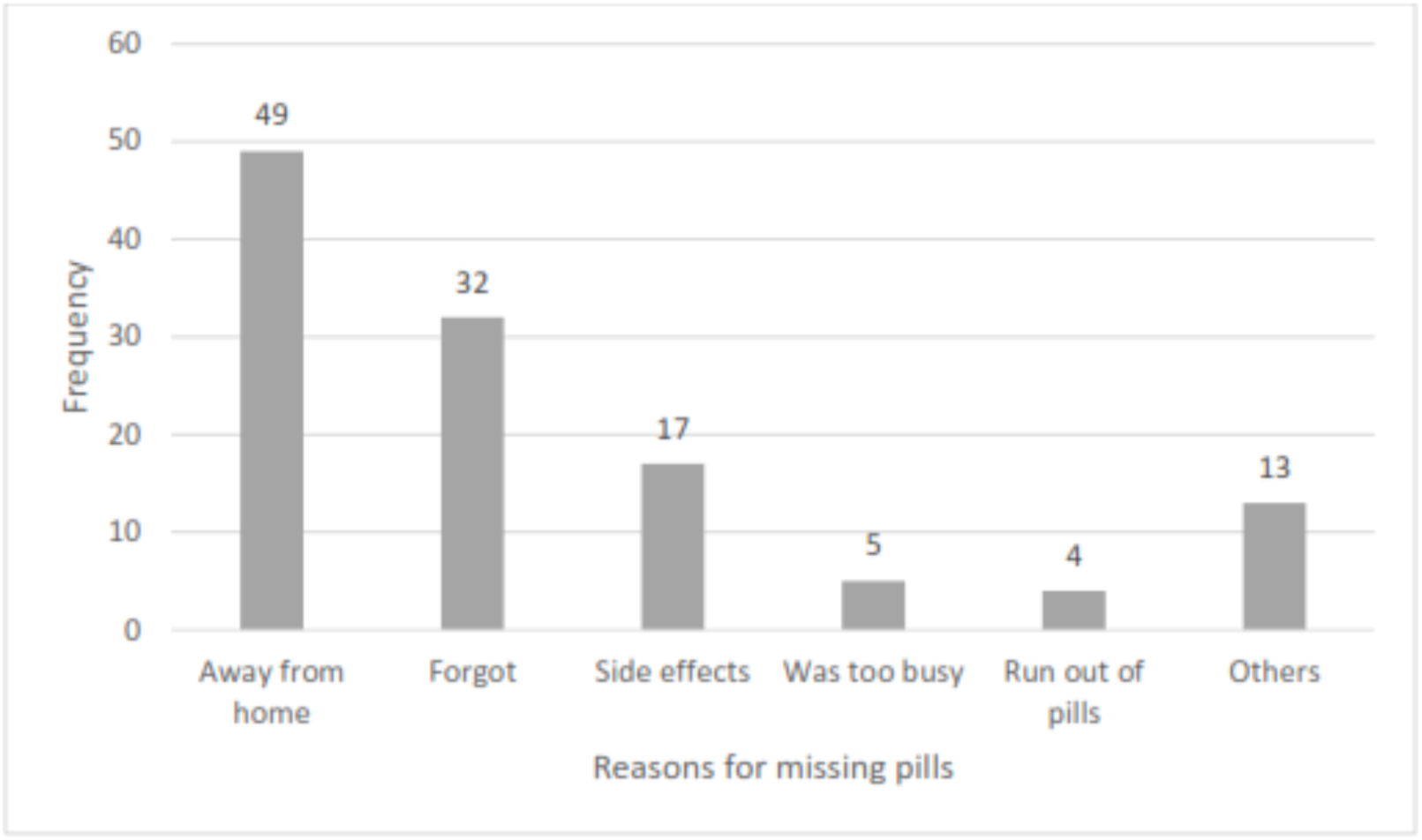
Reasons for missing pills by FSWs (N=120) assessed for adherence, Kampala Uganda 2017.

In multivariable analysis, participants who owned a phone at the time of the study were more likely to adhere to ART than those who did not (AOR 2.90; 95% CI 1.07, 7.88). There was an increased odds of adherence to ART for each ten-year increase in age (AOR 1.60; 95% CI 1.00, 2.60). Compared to married women, widowed participants were less likely to adhere to ART (AOR 0.22; 95% CI 0.05, 0.87). Of note, alcohol use in the last 3 months was not associated with adherence to ART (AOR 0.77; 95% CI 0.42, 1.39).

## Discussion

### Key findings

Our study discovered that adherence to ART was suboptimal among FSW in the GHWP, with only 3 in 5 attaining ≥95% level. Secondly, adherence to ART was independently associated with ownership of a mobile phone, older age by 10–year bands and marital status. Specifically, widows were less adherent to ART. Alcohol use was not associated with adherence in this key population.

### Strengths and limitations

Although our study had some strengths, these findings are better understood with regard to its limitations. We had a relatively large sample size of FSW and conducted this study in a typical urban slum setting for FSW providing for generalisable findings. However, selection bias could have occurred as we enrolled only those FSW who came to the clinic for their visits. Given that As such we possibly over-estimated adherence to ART. Secondly, our measurement of adherence to ART was based on patients’ self-reports of missed pills subject to recall and social desirability biases resulting in overestimated adherence. Good enough, self-reported adherence correlates with viral load changes, is cheaper and more practical in low-income settings [28–30]. Finally, we opted for the most recent viral load results instead of real time viral load tests that could have overestimated adherence.

### Interpretation in relation to existing evidence

Adherence among FSW was relatively low compared to findings from a systematic review of studies largely from higher income countries at 76% [26]. Evidence from other studies reported FSW adherence to ART at 67% [31] and 81% [32]. Additionally, our study reported lower adherence to ART compared to the general populations in sub–Saharan Africa [18, 33]. Low proportions of ART adherence among FSWs have also been reported in India, in a study that estimated adherence at 48% using a combined measure of pill counts and self-reports [25].

The major reason for missing pills by participants in our study was being away from home. Most FSW do not usually operate from their areas of residence where they are known but rather go to other hot spots within the city where they are less likely to be known. It is possible that sometimes they either forget carrying their pills with them or spend more days away from home than had anticipated.

We found a strong association between owning a mobile phone and adherence. This is consistent with that fact most of study participants that reported owning a mobile phone used the phone alarm or clocks as their medicine reminder tool. This is also consistent with findings from a qualitative study conducted in Lesotho that showed that mobile phone alarms and clocks are major facilitators of adherence among people living with HIV [34]. Our findings build a case for the use of mobile phone technologies to enhance adherence to ART as evidenced by the various studies [35–41]. Our study also showed that older age was associated with ART adherence, a finding consistent with what has been found among the general population [18, 33, 42, 43] and other key populations [44, 45]. Though not examined in this study, it is possible that older FSW have lived with HIV for a longer time and therefore more ART experienced than their younger counterparts. This finding suggests the need for targeted interventions to improve adherence among the young FSW. Studies have shown that younger FSW have unequal footing when negotiating in their communities. They are usually street based, poorly educated, occupying low skill and poor paying jobs and always in conflict with their older counterparts due to competition for clients [46, 47]

Additionally, our study found lower odds of adherence to ART among FSW who reported to be widowed. This finding is consistent with findings from a study among the general population of people living with HIV in Dakar, Senegal [48]. Being widowed and a single mother who takes care of the children without a hand of the husband (many times originally the sole bread winner), has been shown to be associated with depression [49]. This, coupled with living in settings where FWS are discriminated and criminalised could possibly affect adherence to ART.

Contrary to our expectation and evidence from other studies [16, 50, 51], data from our study did not show an association between alcohol use and adherence to ART. However, our finding is similar to that from a study among FSWs India [25]. Over half of the FSW in our study reported alcohol use, similar to findings among FSW in another study in Malawi [52] and one in Uganda [6]. Studies have documented use of alcohol and other substances by FSWs as a way of coping with demands of their work [53, 54]. Our measurement of alcohol use was based on a yes/no response to alcohol use in the last three months and not on the standard tool like audit score or cage. This could party explain this contradictory finding. Nonetheless alcohol risk reduction counselling is necessary intervention to FSW. Studies have indicated that alcohol consumption is not only associated with high risk sexual behaviour [6, 53] but may also influence survival of HIV infected patients [55] without altering the effectiveness of ART [56].

### Generalisability

#### Conclusion and recommendations

We found a relatively low proportion of FSW achieving the desirable level of adherence to ART. Our findings build a case for the development and scale up of targeted intervention strategies to increase to ART adherence among FSWs. Special consideration to young FSW is highly recommended based on the findings of our study. Incorporation of mobile phone technology applications in the routine adherence counselling follow-up services should be explored further and embraced as appropriate. These could include adherence counselling text message reminders, voice notes or even peer support information technology platform especially to the young FSWs.

## Data Availability

All data produced in the present study are available upon reasonable request to the authors

